# Support mechanisms for nurses aspiring to or undertaking clinical academic development globally – protocol for a scoping review

**DOI:** 10.1101/2023.08.06.23293389

**Authors:** Claire Jennings, Gill Norman, Marie Marshall, Michelle Briggs

## Abstract

**Introduction:** Nurse researchers often lack awareness of how to start a clinical academic research career and often lack clear entry routes. This scoping review aims to identify the range and nature of clinical academic opportunities that are available for nurses. This will identify knowledge gaps and provide the basis for future research.

**Methods and analysis:** The review will be conducted using scoping review methodology and reported in accordance with PRISMA-SCR guidelines. We will search CINAHL (EBSCO), Medline (OVID), AMED (OVID) and ProQuest using a search based on the facet’s ‘nurse’ ‘research’ and ‘clinical academic’. Grey literature and a hand search of the reference lists will be conducted for additional publications which meet the inclusion and exclusion criteria. We will include articles with a focus on nurses who are interested in undertaking clinical academic development, or a focus on programmes aimed at supporting clinical academic development for nurses. Following initial screening of titles and abstracts, relevant full text papers will be screened for inclusion. A proportion of all records at each stage will be screened by a second reviewer. We will use a prespecified form to extract data which will be charted and presented in tabular form. Samples of data extraction and charting will be checked by a second reviewer. This will support a narrative summary structured around key identified variables agreed iteratively by the review team.

**Ethics and dissemination:** No ethical or Health Research Authority approval is required to undertake this scoping review. This protocol has been registered with The Open Science Framework (DOI 10.17605/OSF.IO/WVDXH). The findings will inform future research exploring the support mechanisms for nurses aspiring to undertake clinical academic development and be disseminated via presentations at national conferences and publications in peer reviewed journals.

**Article summary:** *Strengths and Limitations of this study:*

- This is the first exploration review of the nature and range of clinical academic opportunities that are available to nurses.
- The scoping review will be undertaken and reported following established guidelines to ensure transparency and reproducibility.
- The identification and synthesis of data will include published articles found on the CINAHL, Medline, AMED and ProQuest databases, grey literature available on relevant organisations and professional bodies, and a hand search of reference lists.
- The scoping review will be focussed on nurses undertaking clinical academic development programmes, as they are the largest group of registered healthcare professionals and are underrepresented as research leaders.
- Due to limited resources, only a proportion of the screening will be undertaken be 2^nd^ reviewer, and literature not available in English Language will be excluded. We will mitigate these issues by use of additional checks and listing of relevant foreign language studies.

## Background/Rationale

Clinical academics in nursing, midwifery and allied health professions (NMAHP) play a pivotal role within the NHS by bringing a patient-focused insight to the fore and conducting translational research which offers direct benefits to the quality of patient care (Council of Deans, 2013).

There are key differences between the nursing workforce versus midwifery and allied health professional’s workforce is in terms of size and complexity, nurses have been the largest clinical workforce in the UK since 1948 (NHS, 2023). The last six years have seen publication of focussed research strategies for nursing, midwifery, and allied health professions separately (NHS England, 2021; Scottish Government, 2017; Welsh Government, 2022). This has given impetus to the need to explore research led by nurses, and the contributions they can make as members of multidisciplinary research teams to drive change.

The National Institute for Health Research (NIHR) has recognised that nurses and midwives are underrepresented, and in 2019 the NIHR Nursing and Midwifery Incubator was established to accelerate capacity building and support the development of a skilled clinical academic research workforce across the nursing and midwifery professions (NIHR, 2022).

However, despite efforts to highlight clinical academic careers, the number of nurses developing a profile is disproportionately low relative to all health professionals applying for clinical doctoral fellowships (Gibson, 2019). Nurses often lack awareness of how to start a clinical academic research career and clear and well-publicised routes of entry are rare. Subsequently the career structure and guidance about how to navigate of combining a clinical role with academic development is not well established. Meanwhile the many different development opportunities becoming available to support nurses embarking on clinical academic careers, the absence of an established structure may cause difficulties for clinical managers when trying to plan their operational service.

In addition to formal routes, mentoring, coaching and advice can be a necessary step to help professional and personal development and aspirations in early career clinical academics (Baltruks and Callaghan, 2018). This support process can help clinical academics by providing pastoral care and help in dealing with the demands of a clinical academic research career (Baltruks and Callaghan, 2018). However, it is unclear what information is available in literature about the support enabling nurses to develop as clinical academics.

For these reasons, a scoping review will be conducted to systematically map the research done in this area, as well as to identify any existing gaps in knowledge. (Arksey and O’Malley, 2005).

Scoping reviews have shown to be an essential tool to summarise all studies, in an accessible document, that could inform future actions (Dobrowolska et al., 2021; Vassie et al., 2020).

## Objective

The objective of this scoping review is to systematically scope the literature to identify the range and nature of clinical academic opportunities that are available or can be accessed by nurses. It will also clarify and map key concepts and definitions underpinning the supervision required for nurses undertaking clinical academic development, the metrics reported for implementation and outputs from a clinical academic programme or pathway.

## Methods

The scoping review will be guided by the methodological frameworks proposed by Arksey and O’Malley (2005) and the Joanna Briggs Institute (Tricco et al., 2018). Thus, the following five steps will be followed in this scoping review:

1. Identifying the research question,
2. Identifying relevant studies,
3. Selection of eligible studies,
4. Charting the data,
5. Collating and summarizing the results.

This review will follow the relevant aspects of the Preferred Reporting Items for Systematics Review with the Extension for Scoping Reviews (PRISMA-ScR) to ensure thorough reporting and mapping of the body of literature (Khalil et al., 2021).

## Identifying the research question

The ‘PCC’ approach uses definition of the population, concept and context to support a clear and meaningful title, objective and inclusion criteria for a scoping review (Peters et al., 2020)).

### Population/types of participants

this review will consider all articles that relate to registered nurses. Nursing is a profession which is regulated by law in the UK, and the Professional Qualifications Bill (UK Parliament, 2022) sets out that a profession is regulated where there is a legal requirement to have certain qualifications or experience to undertake certain professional activities or use a protected title (Department for Business Energy & Industrial Strategy, 2021).

### Concept

The focus will be on nurses who are aspiring to explore or pursue clinical academic development, or who already identify as a clinical academic. A clinical academic will be defined as engaging concurrently in clinical practice and research, or provide clinical and research leadership in the pursuit of innovation, scholarship and provision of excellent evidence-based healthcare (Department of Health, 2012) As part of this the review we will also consider articles that illustrate, investigate or explore programmes or courses that support clinical academic development and are available to nurses, such as internships, fellowships, or doctorates.

### Context

The context of this review is any healthcare system in high income countries (World Bank Group, 2022), including acute care, primary care, social care, rehabilitation, and community settings.

The main research question will be ‘What information is available about the clinical academic opportunities that are available to nurses that are aspiring to or pursuing early clinical academic career research development?

The research sub-questions are as follows:

- What are the aims and characteristics (type, duration, provider) of clinical academic development programmes?
- What is the uptake and accessibility of the programmes on offer?
- What constitutes success (metrics, output) for nurses on aspiring to or pursuing clinical academic development?
- What support (mentorship, supervision, funding) is available for nurses undertaking the programmes on offer?
- What are the experiences of those undertaking such development, and those that are supporting and supervising on such programmes

## Identifying relevant studies

A systematic search strategy was developed to identify the relevant studies for inclusion in the review. Recognition of the need to develop nurses as clinical academics, along with midwives and allied health professionals, has become prominent in the last 20 years, however we will not use date limits for the search to ensure inclusion of any earlier seminal work in this field.

The following electronic databases will be searched

- Cumulative Index to Nursing and Allied Health Literature (CINAHL) (Ebsco)
- MEDLINE (Ovid)
- AMED (Ovid)
- PROQUEST

An expert university librarian assisted the development of the search terms to ensure that all relevant material was identified. Terms are searched as both keywords in the title and/or abstract and subject headings (e.g., MeSH, EMTREE) as appropriate. The full search strategies for each database are given in the supplementary material.

We will search grey literature. This will include searching websites of relevant organisations or professional bodies for any documents related to clinical academic development or programmes. Organisations have been identified from a preliminary search of the internet via Google, such as the National Institute of Health Research, and Health Education England

Reference lists of included studies will be checked for further studies.

## Selection of eligible studies

Title and abstract screening were guided by the PCC framework (Table 1). We will include records which meet the following inclusion criteria:

- A focus on, or including a majority of, nurses who are interested in pursuing or are already undertaking clinical academic development or a focus on programmes aimed at encouraging, supporting, or advancing clinical academic careers or development for nurses. Depending on the volume of literature identified we may limit inclusion to studies where the focus is nurses or at least 50% of participants are nurses.
- Available in English language (due to lack of funding for translation services)

We will exclude records with any of the following:

- Text that does not refer to nurses interested in or exploring clinical academic development
- Full-text articles that could not be obtained or full text that is not in available in English language,
- Studies available in abstract only

All identified records will be uploaded into Endnote online software (Clarivate, 2023) for deduplication. Records will then be uploaded to Rayyan software (Ouzzani et al., 2016) for screening of both title and abstract, and of full text. This supports blinded screening of a proportion of records and shared decision making between the reviewers.

Abstracts will be reviewed for relevance to the review question and where potentially eligible the full text will be obtained. Following initial piloting on a small subset of records, one reviewer (CJ) will screen the records and at each stage a minimum of 10% of records will be independently screened by a second reviewer (GN). Disagreements will be resolved through consensus and if necessary, consultation with a third reviewer (MB). A second reviewer will also be consulted in cases of uncertainty. This approach will also be used for full text records. Records excluded at full text will be listed in the review with reasons for exclusion, to ensure transparency and rigour. The screening process will be documented in a PRISMA flow diagram.

## Charting the data

We will develop a bespoke data extraction form in Microsoft Excel, based on scoping review guidance (Peters et al., 2020). This data extraction tool will be piloted by two reviewers using a small number of papers to ensure that the data relevant to the research question will be collected fully from the articles identified.

The data extraction form will include details of the author(s), year of publication, population studied, context, concept, methodology used and key findings relevant to nurses interested or undertaking clinical academic development. A separate tool may be used for policy documents or guidance if appropriate. Extraction will be undertaken by one reviewer (CJ) and a second reviewer (GN) will check a proportion of the data extraction, to ensure a standardised process; a second reviewer will also be conducted in cases of uncertainty. Any disagreements will be resolved through consensus or by consultation with a third reviewer (MM or MB).

We anticipate that there may be a need for iterative development of the data extraction approach. Any amendments to the tool after piloting is complete will be recorded including the reasons for this, for transparency. The final version of the data extraction tool will be included in the appendix of the scoping review report to ensure reproducibility.

## Collating and summarising the results

The extracted data will be charted to map key characteristics and then represented in tabular form and as a narrative summary.

The tables will report the following:

- Publication/production year and country of origin
- Programme design and aims/objectives
- Professional background of nurses that are the focus for such programmes
- Characteristics of clinical academic development, such as:
  ∘ Marketing and dissemination of programme
  ∘ Application requirements
  ∘ Selection process
  ∘ Structure
  ∘ Duration
  ∘ Provider
  ∘ Funding
  ∘ Educational strategies and methods employed within the programme
  Assessment within the programme
- Outcomes or metrics reported including participant or provider experience
- Supervision, support, and mentorship offered or provided to support those interested in or undertaking clinical academic development.

The narrative summary will be structured by the different types of training and programmes available and at whom they are aimed. Additionally, it will focus on different types of formal and informal mechanisms used to support nurses undertaking clinical academic careers. It will also consider the role of support from managers, mentors, or coaches.

## Ethical Considerations

No ethical or Health Research Authority approval is required to undertake this scoping review. The protocol has been registered with the OSF (OSF, 2011) to ensure quality through transparent reporting and to prevent overlapping or duplicate work being undertaken prior to publication of the review findings (https://doi.org/10.17605/OSF.IO/ACNZP)

## Dissemination

The scoping review will also be disseminated by publication in a peer reviewed journal (preprint may be used on submission) and submitted for presentation at a national conference. We may also use professional networks to disseminate the results to a wider audience of nurses, midwives and allied healthcare professionals undertaking clinical academic development.

## Patient and public involvement

We did not undertake any public or patient involvement in developing this protocol. This is because the review maps out evidence intended for research capacity building of the nursing workforce.

## Discussion

The findings of this scoping review will inform future work with nurses to identify their awareness and experience of clinical academic careers, and to explore the lived experiences nurse researchers, understanding the pathways they have undertaken and the support networks they have utilised.

This information could be used by decision-makers to develop proactive plans to support the development of programmes which aim to support nurses pursuing clinical academic development. The findings could also be drawn on when planning curriculum and continued professional development activities for non-medical healthcare professionals inspired to pursue a clinical academic career.

## Data Availability

This scoping review protocol has been registered with The Open Science Framework (DOI 10.17605/OSF.IO/WVDXH). All data generated through the review process will either be made available as supplementary material in publication of the review or will be available on reasonable request to the authors.

## Author contributions

CJ had the idea for the review, devised the scoping review question, methodology and wrote the first draft of the protocol. GN, MB and MM substantively contributed to the development of the scoping review question and methodology and revised and approved the protocol.

## Acknowledgements

We are grateful to the librarians at the University of Manchester for their assistance with the process of refining the search strategy.

We are also grateful to the librarians at Manchester University NHS Foundation Trust for their support in locating full-text articles.

## Competing interest statements

All authors have completed the ICMJE uniform disclosure form at http://www.icmje.org/disclosure-of-interest/ and declare:

GN supervises allied health professionals - including nurses - as interns, predoctoral fellows and PhD students for the University of Manchester and the NIHR Applied Research Collaboration – Greater Manchester.

MM is the Deputy Director of Nursing for NMAHP Research & Innovation at Royal Manchester Children’s Hospital in Manchester University NHS Foundation Trust, and Clinical Lead for the Women & Children Domain of the Manchester Academic Health Science Centre.

MB is the Deputy Chair of UK Council of Deans Clinical Academic Careers Implementation Network, steering group member of NIHR Nursing and Midwifery Incubator. Academic Career Development Lead for NIHR Applied Research Collaboration (ARC_GM), Director of the Manchester Clinical Academic Centre for Nurses, Midwives and AHPs and funding panel member for NIHR Doctoral Clinical and Practitioner Academic Fellowship Programme.

