## Supplemental file for "Support mechanisms for nurses aspiring to or undertaking clinical academic development globally – protocol for a scoping review"

**Scoping Review Protocol – Supplementary material**

**Search strategy**

CINAHL

| S1 | TI Nurs* OR AB Nurs* |
| --- | --- |
| S2 | TI ‘Clinical academi*’ OR AB ‘Clinical academi*’ |
| S3 | TI ‘Research career*’ OR AB ‘Research career*’ |
| S4 | TI ‘Research internship*’ OR AB ‘Research internship*’ |
| S5 | (MM "Nurse Researchers") |
| S6 | (MM "Students, Nursing, Doctoral") |
| S7 | "Nurse Researcher" |
| S8 | "Students, Nursing, Doctoral" |
| S9 | S2 OR S3 OR S4 OR S5 OR S6 OR S7 OR S8 |
| S10 | S1 AND S9 |

Medline

| 1 | Nurs*.ti. or Nurs*.ab. or Nurs*.kw. |
| --- | --- |
| 2 | 'Clinical academi*'.ti. or 'Clinical academi*'.ab. or 'Clinical academi*'.kw. |
| 3 | 'Research* training'.ti. or 'Research* training'.ab. or 'Research* training'.kw. |
| 4 | 'Research intern*'.ti. or 'Research intern*'.ab. or 'Research intern*'.kw. |
| 5 | 'Research career*'.ti. or 'Research career*'.ab. or 'Research career*'.kw. |
| 6 | *Nursing Methodology Research/ed, mt, og [Education, Methods, Organization & Administration] |
| 7 | *Clinical Nursing Research/ed, mt, og [Education, Methods, Organization & Administration] |
| 8 | Nurse researcher.mp. |
| 9 | 2 or 3 or 4 or 5 or 6 or 7 or 8 |
| 10 | 1 and 9 |

AMED

| 1 | Nurs*.ti. or Nurs*.ab. or Nurs*.sh. |
| --- | --- |
| 2 | 'Clinical academi*'.ti. or "Clinical academi*".ab. or "Clinical academi*".sh. |
| 3 | 'Research* training'.ti. or 'Research* training'.ab. or 'Research* training'.sh. |
| 4 | 'Research intern*'.ti. or 'Research intern*'.ab. or 'Research intern*'.sh. |
| 5 | 'Research career*'.ti. or 'Research career*'.ab. or 'Research career*'.sh. |
| 6 | Fellowship*.ti. or Fellowship*.ab. or Fellowship*.sh. |
| 7 | Nurse researcher.mp. |
| 8 | Student nursing doctoral.mp. |
| 9 | 2 or 3 or 4 or 5 or 6 or 7 or 8 |
| 10 | 1 and 9 |

ProQUEST

| S1 | (((title(Nurs*) OR abstract(Nurs*)) AND stype.exact("Conference Papers & Proceedings" OR "Government & Official Publications" OR "Reports" OR "Working Papers" OR "Scholarly Journals" OR "Dissertations & Theses")) AND at.exact("Report" OR "Dissertation/Thesis" OR "Government & Official Document" OR "Working Paper/Pre-Print" OR "Literature Review" OR "Reference Document" OR "Review" OR "Conference Proceeding" OR "Conference Paper" OR "Conference" OR "Article")) AND la.exact("English") |
| --- | --- |
| S2 | (((title("Clinical academi*") OR abstract("Clinical academi*")) AND stype.exact("Conference Papers & Proceedings" OR "Government & Official Publications" OR "Reports" OR "Working Papers" OR "Scholarly Journals" OR "Dissertations & Theses")) AND at.exact("Report" OR "Dissertation/Thesis" OR "Government & Official Document" OR "Working Paper/Pre-Print" OR "Literature Review" OR "Reference Document" OR "Review" OR "Conference "Review" OR "Conference Proceeding" OR "Conference Paper" OR "Conference" OR "Article")) AND la.exact("English") |
| S3 | (((title(“Research* training”) OR abstract(“Research* training”)) AND stype.exact("Conference Papers & Proceedings" OR "Government & Official Publications" OR "Reports" OR "Working Papers" OR "Scholarly Journals" OR "Dissertations & Theses")) AND at.exact("Report" OR "Dissertation/Thesis" OR "Government & Official Document" OR "Working Paper/Pre-Print" OR "Literature Review" OR "Reference Document" OR "Review" OR "Conference Proceeding" OR "Conference Paper" OR "Conference" OR "Article")) AND la.exact("English") |
| S4 | (((title(“Research intern*”) OR abstract(“Research intern*”)) AND stype.exact("Conference Papers & Proceedings" OR "Government & Official Publications" OR "Reports" OR "Working Papers" OR "Scholarly Journals" OR "Dissertations & Theses")) AND at.exact("Report" OR "Dissertation/Thesis" OR "Government & Official Document" OR "Working Paper/Pre-Print" OR "Literature Review" OR "Reference Document" OR "Review" OR "Conference Proceeding" OR "Conference Paper" OR "Conference" OR "Article")) AND la.exact("English") |
| S5 | (((title(“Research career*”) OR abstract(“Research career*”)) AND stype.exact("Conference Papers & Proceedings" OR "Government & Official Publications" OR "Reports" OR "Working Papers" OR "Scholarly Journals" OR "Dissertations & Theses")) AND at.exact("Report" OR "Dissertation/Thesis" OR "Government & Official Document" OR "Working Paper/Pre-Print" OR "Literature Review" OR "Reference Document" OR "Review" OR "Conference Proceeding" OR "Conference Paper" OR "Conference" OR "Article")) AND la.exact("English") |
| S6 | (((title(“Fellowship*”) OR abstract(“Fellowship*”)) AND stype.exact("Conference Papers & Proceedings" OR "Government & Official Publications" OR "Reports" OR "Working Papers" OR "Scholarly Journals" OR "Dissertations & Theses")) AND at.exact("Report" OR "Dissertation/Thesis" OR "Government & Official Document" OR "Working Paper/Pre-Print" OR "Literature Review" OR "Reference Document" OR "Review" OR "Conference Proceeding" OR "Conference Paper" OR "Conference" OR "Article")) AND la.exact("English") |
| S7 | S2 OR S3 OR S4 OR S5 OR S6 |
| S8 | S1 AND S7 |
